# Financial conflicts of interest between pharmaceutical companies and executive board members of internal medicine subspecialty societies in Japan between 2016 and 2020

**DOI:** 10.1101/2023.04.18.22282273

**Authors:** Anju Murayama, Hiroaki Saito, Tetsuya Tanimoto, Akihiko Ozaki

## Abstract

Using payment data disclosed by 92 major pharmaceutical companies, this study evaluated personal payments to executive board members (EBMs) among each specialized field of internal medicine in Japan. A total of 63,405 personal payments worth of $70,796,014 in payment amounts were made to 99.2% of all EBMs of the 15 leading professional societies of internal medicine in Japan between 2016 and 2020.

## Background

Pharmaceutical companies make payments to physicians for several purposes such as lecturing or consulting compensations. Such payments can improperly bias physicians’ clinical practice to the possible detriment of patients. This is particularly concerning in Japan, where the 2016 annual turnover of pharmaceutical products was as high as $88.0 billion.

To help improve transparency concerning the financial conflicts of interest (COIs) of physicians in Japan, each company affiliated with the Japan Pharmaceutical Manufacturers Association (JPMA) has been required to disclose on their website all annual payment data to individual physicians since 2013. Using this data, we previously reported that, among the executive board members (EBMs) of the Japanese professional medical associations, the specialty of internal medicine attracted more payments than others(1).

However, as differences among sub-specialties of internal medicine were unknown, we analyzed payments to EBMs among each specialized field of internal medicine to identify any such differences.

## Methods

This cross-sectional study evaluated the financial relationships between pharmaceutical companies and the EBMs of 15 medical associations representing different sub-specialties determined by the Japanese Society of Internal Medicine. We considered all EBMs who were positioned in 2019 and collected all personal payment data made to the EBMs in the current year (2019), three years before (2016-2018) and one year after (2020) their board membership. Personal payments for lecturing, writing, and consulting work were collected from a total of 92 pharmaceutical companies belonging to the JPMA between 2016 and 2020, as we reported previously(2). Then, descriptive analyses were conducted. Additionally, we reviewed the COIs policies of each society and the disclosure status of the EMBs’ COIs. This study was approved by the Ethics Committee of the Medical Governance Research Institute.

## Results

Of the 353 different EBMs identified, 350 (99.2%) received one or more personal payments from the pharmaceutical companies over the five years. 99.2% (350) and 97.2% (343) of all EBMs received personal payments three years before and in the year of their board membership. A total of 63,405 personal payments worth of $70,796,014 in payment amounts were made to the EBMs over the five years. 81.1% ($57,444,620) of total payments were made for lecturing fees, which are typically compensations for speaking about novel drugs at event sponsored by the pharmaceutical companies. The median five-year combined personal payments per EBM was $150,849 (interquartile range [IQR]: $73,412 - $282,456). EBMs who were chairman or vice chairman of executive board received significantly larger median personal payments than those who were not ($225,685 vs $143,885, p=0.01 in the U test).

Among the 15 societies, there were 12 (80.0%) societies with all EBMs receiving personal payments from the pharmaceutical companies. The EBMs of the Japan Diabetes Society received the highest median payments of $306,041 per EBM, followed by the Japanese Society of Hematology (median: $212,703), and Japanese Society of Allergology (median: $210,316). (Figure 1) Although every society has their own conflicts of interest policy, none publicly discloses the financial relationships between pharmaceutical companies and their EBMs due to their privacy.

**Figure 1.**
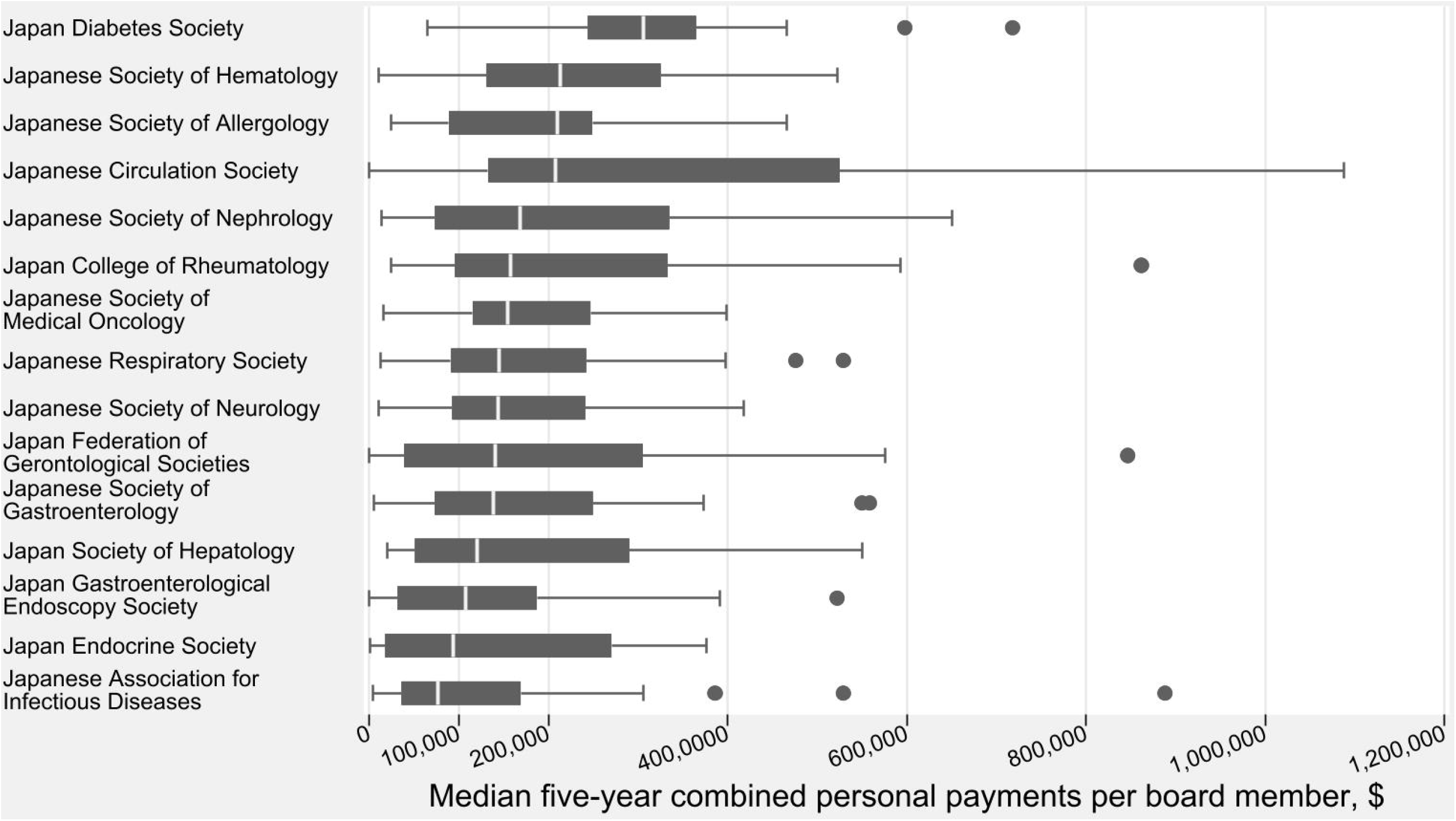
Median personal payments from pharmaceutical companies to the society board members between 2016 and 2020 Japanese yen were converted to US dollars using the 2020 average monthly exchange rate (106.77 JPY per 1 USD).

## Discussion

Despite several limitations including possible inaccuracies in the payment information and the limited types of payments, this analysis of payment information publicly disclosed by the pharmaceutical companies demonstrated that, nearly all (99.2%) EBMs of the 15 leading professional societies of internal medicine in Japan received more than $70 million in personal compensations in the past five years. Additionally, these financial relationships were not publicly disclosed on the society webpages.

The substantial financial relationships between EBMs and healthcare industry were previously documented in the US(3), France(4), and Australia(5). However, any of the studies showed that smaller amounts of payments were made to lower fraction of EBMs than those in Japan. Seventy-two percent of the US EBMs received $31,805 in median over six years(3). The higher percentage of influential physicians receiving large personal fees in Japan than in other countries was consistent with previous research on clinical practice guideline authors in Japan(2,6). Additionally, the COI disclosures of the EBMs was not fit for purpose. Given that we previously reported a similar unsatisfactory status concerning physicians, including EBMs and clinical practice guideline authors in Japan(1), an improved system for COI disclosure is imperative. Transparency concerning COIs of EBMs should be improved to ease the concern of patients as well as gain more trust among the general public in Japan.

## Data Availability

All data produced in the present study are available upon reasonable request to the authors.

## Sources of financial support that require acknowledgement

The Medical Governance Research Institution and the Tansa provided financial support for the present research. Tansa is a non-profit organization engaged in objective journalism and is a member of the Global Investigative Journalism Network. The Medical Governance Research Institution is a non-profit organization and has received donations from various individuals, industries, and organizations, including donations from Ain Pharmacies, Japan. The funders were not involved in the design of the study, the work carried out, or interpretation of the study findings.

## Acknowledgement

The authors appreciate Dr. Masahiro Kami, M.D., PhD (Medical Governance Research Institute) for his constructive opinion, Ms. Erika Yamashita (Medical Governance Research Institute) for her data cleaning, Ms. Tomoyo Nishimura for her persistent support for the present study, the Tansa for acquisition of the payment data, and Editage (www.editage.jp) for commercial English language editing.

## Abbreviation

JPMA: Japan Pharmaceutical Manufacturers Association
EBM: executive board members

